# Accurate influenza forecasts using type-specific incidence data for small geographical units

**DOI:** 10.1101/19012807

**Authors:** James Turtle, Pete Riley, Michal Ben-Nun, Steven Riley

## Abstract

Influenza incidence forecasting is used to facilitate better health system planning and could potentially be used to allow at-risk individuals to modify their behavior. For example, the US Centers for Disease Control and Prevention (CDC) runs an annual competition to forecast influenza-like illness (ILI) at the regional and national levels in the US, based on a standard discretized incidence scale. Here, we use a suite of forecasting models to analyze type-specific incidence at the smaller spatial scale of clusters of nearby counties. We used data from point-of-care (POC) diagnostic machines over three seasons, in 10 clusters, capturing: 57 counties; 1,061,891 total specimens; and 173,909 specimens positive for Influenza A. Total specimens were closely correlated with comparable CDC ILI data. Mechanistic models were substantially more accurate when forecasting influenza A positive POC data than total specimen POC data, especially at longer lead times. Also, models that fit individual counties separately were better able to forecast clusters than were models that directly fit to aggregated cluster data. Public health authorities may wish to consider developing forecasting pipelines for type-specific POC data in addition to ILI data. Simple mechanistic models will likely improve forecast accuracy when applied at small spatial scales to pathogen-specific data before being scaled to larger geographical units and broader syndromic data. Highly local forecasts may enable new public health messaging to encourage at-risk individuals to temporarily reduce their social mixing.

## Introduction

Influenza infections cause substantial morbidity and mortality across all geographical areas and sociodemographic groups (1). Forecasting pipelines -- of data and associated analytics -- can create knowledge of current and likely future incidence and therefore have high potential value to individuals and health planners (2), potentially leading to at-risk individuals choosing to modify their behaviour to reduce their individual risk when incidence is high (3). Also, health systems could more efficiently manage scarce resources such as intensive care services that are impacted by local influenza incidence but also utilized by many different healthcare pathways (4). More generally, seasonal influenza is an ideal case study for influenza pandemics, with forecasting during influenza pandemics and other global health outbreaks is a key priority to support rapid investment decisions and optimization of available resources (5, 6).

Current forecasting pipelines for influenza have focused on the use of syndromic data for large geographically diffuse populations (7). For example, the US Centers for Disease Control and Prevention (CDC) forecasting challenge produces public forecasts of influenza-like illness rates at regional and national scales for the US, based on a long-running national syndromic surveillance system that has proven robust during a pandemic (8). Participating teams use a variety of methods. Ensembles of fully automated models (2) have been shown to improve average performance over individual statistical (9) and mechanistic models (10) but do not yet outperform crowd-sourced expert opinion (11) for key forecasting targets (7).

Here, we consider the possibility that alternate data streams may be much more closely related to the underlying biology and hence be inherently more forecastable using mechanistic models (10). Specifically, that more local and pathogen-specific data may permit more accurate forecasting pipelines with mechanistic models than are currently possible using more geographically aggregated syndromic data (7) or low resolution type-specific data cross references with local syndromic proxy data (12). We apply a set of previously validated models (10) to incidence data from geolocated point-of-care diagnostic devices (13), and use a skill scoring system that is analogous to that used thus far (14, 15) for the CDC prospective competition to compare the forecastability of alternate data streams.

## Results

We obtained 3,258,166 specimen results from 12,227 point-of-care (POC) machines in the United States starting from week 27, 2016 through week 26, 2019. These data were down-selected to obtain clusters of nearby counties with at least 250 tested specimens in any season, resulting in 10 clusters, capturing: 57 counties (Table S2); 1,061,891 total specimens; and 173,909 specimens positive for Influenza A (Figure 1, see Methods). A cluster containing metropolitan Atlanta was just below the average season requirements for 2016-17 but had very high coverage for 2017-18 and 2018-19 and was thus included.

**Figure 1.**
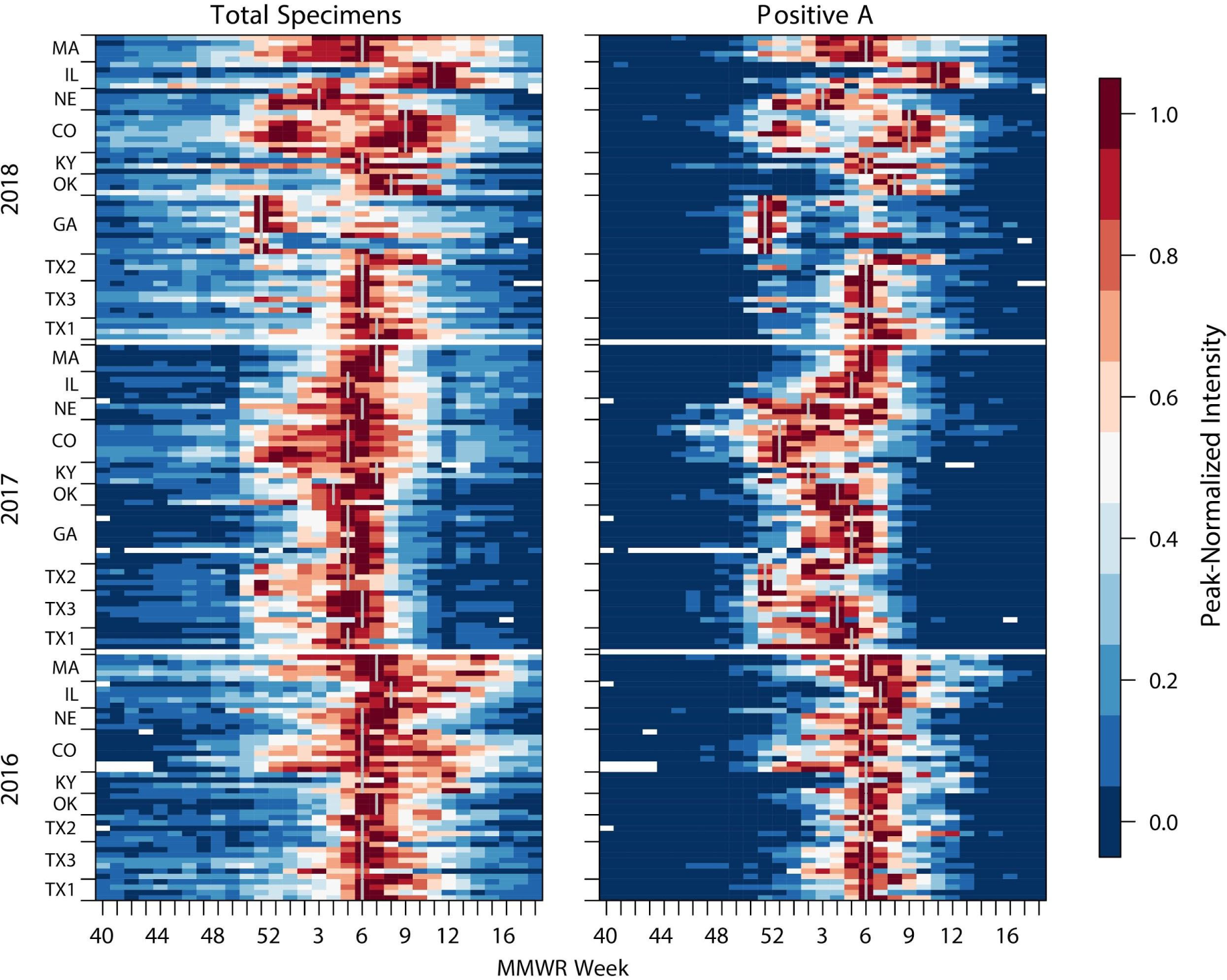
Incidence for each cluster for syndromic (Total Specimens, LHS) and specimens positive for A, RHS). Each row of pixels represents a single county with counties grouped into clusters (bounded by tick marks) and then into seasons. Seasons are indicated by years on the y-axis (with a blank row between years). Within seasons, clusters are ordered by latitude and within clusters, counties are also ordered by latitude. Cluster peak weeks are marked with a gray vertical line. In this figure, the number of reports for each week is normalized for each county and year using the maximum number for that year (Legend, RHS).

Total specimens (TS) and specimens positive for influenza A (PA) both showed broadly similar dynamics, however, there were some intriguing differences between them (Figure 1). As would be expected, the more specific PA data had a clearer difference between off-season and in-season. Also, regardless of noise during periods of low activity, the county-level PA time series produced sharper epidemics, with 6.05 weeks average duration at or above half maximum incidence compared with 8.84 weeks for the TS data. For clusters, the peaks became slightly wider with a width at half peak for PA averaging 6.48 weeks and TS averaging 9.57 weeks. The TS data showed a high correlation with influenza-like illness data for the enclosing CDC region (overall average Pearson coefficient 0.96, Fig S1, Table S1).

When averaging across all clusters, at all forecast weeks, for all years; we found substantial evidence that PA data could be more accurately forecast than could TS data (Figs 2, S2, S3, S4). We assessed accuracy using a similar scoring system as that used for influenza by the US CDC (7). The historical maximum range was separated into 131 bins and we used our model to assign a probability to every possible category. Forecast skill was defined as the sum of probabilities assigned to the eventually observed bin and the 5 immediately lower and higher bins (for a total of 11 bins). Forecast score was the natural log of forecast skill. For example, if we assigned a total probability of 20% to bins in the skill window, then the score would be ln(0.2) = −1.6. The best model for 2-week ahead forecasts scored an average of −1.41 for PA data compared with −2.55 for TS data. Although the accuracy of the forecast did vary over the season, the superiority of best fits to PA data over TS data was maintained across the whole season. In fact, the accuracy of the best fit model for PA data with a 10-week lead time was still better than that of the TS data at a 2-week lead time.

**Figure 2.**
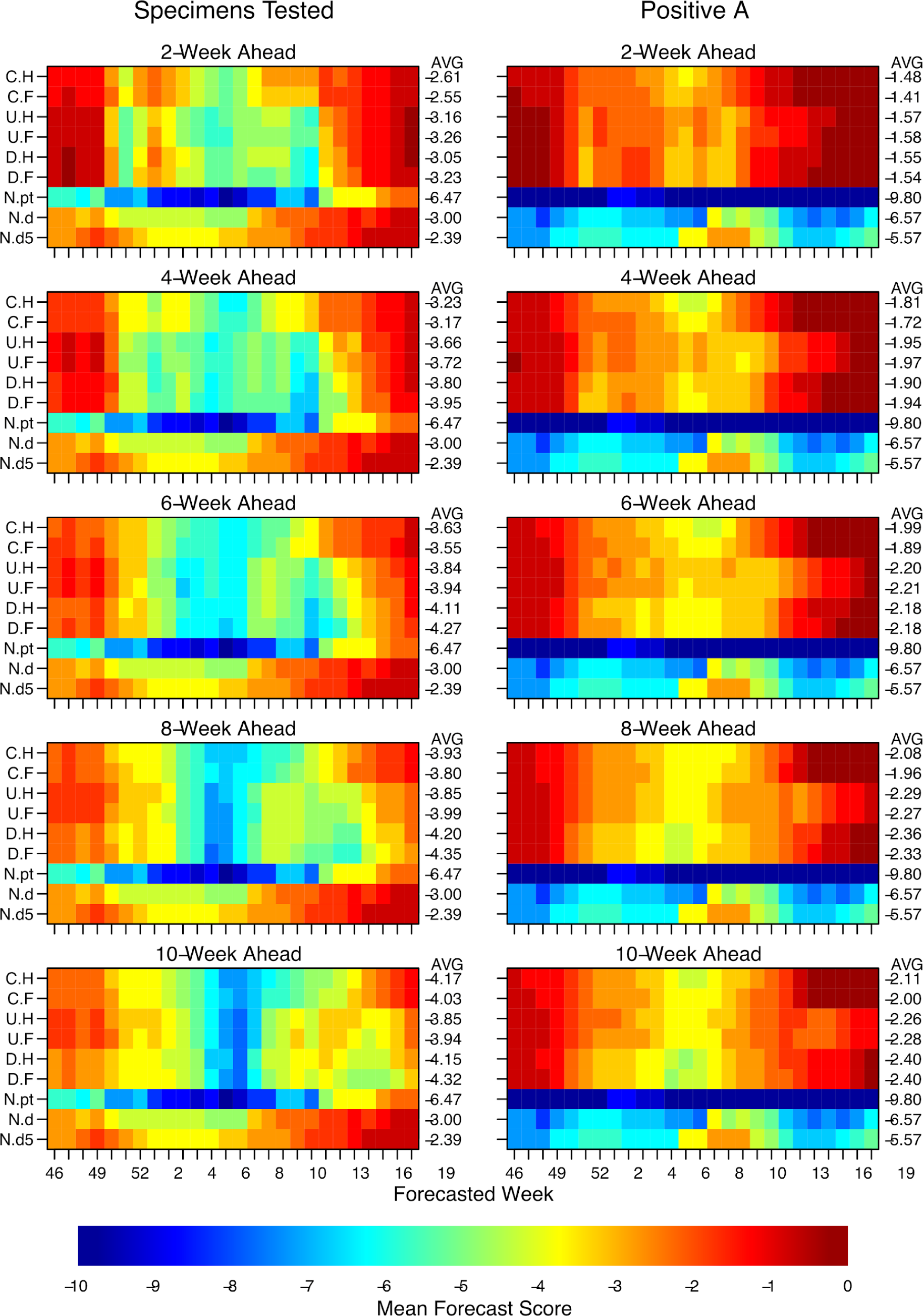
Forecast scores lead times from 2- to 10-weeks ahead, for different specimen types. Pixel colour shows forecast score (see main text) for a given observation week averaged across clusters and seasons, e.g. the 4-week ahead forecast for week 48 was made using data only upto week 44. Averages across all weeks for a given model are printed on the RHS of each row of pixels. Model type is shown on LHS y-axis tick labels: C.H, coupled model with humidity modulated contact rate; C.F, coupled model with fixed contact rate; U.H, uncoupled with humidity; U.F, uncoupled with fixed contact rate; D.H, model directly fitted to cluster with humidity term; D.F, model directly fitted to cluster with fixed contact rate; N.pt, null model made from simple model of that week for all other seasons; N.d, null model made from fitting a log normal to all observations for that week from other years; and N.d5, null model made from fitting log-normal to the observation week, two weeks prior and two weeks following for all other years (see main text). Models are ordered approximately from least complex on the bottom rows to most complex on the top row. See Figures S2 and S3 for complete 1-10 weeks ahead forecast results.

We considered specific examples to understand how and why the models were better able to fit these data in different scenarios. For example, after testing all forecast models at all forecast times for the Colorado cluster for 2016-17, the best fitting model was better able to capture the rapid acceleration of incidence and the shape of the peak PA data stream than was possible for the best fitting model for the TS data (Fig 3 A and B). Despite not being able to accurately fit the decay phase of the epidemic, the best fitting model for the PA data still achieved a substantially better score of −0.51 than the score of −0.82 that was possible for the corresponding TS data. We note that a score of −0.51 corresponds to 60% of forecasts falling within the skill window. However, for the same cluster for the following year, the same did not apply. The best model fits to both datasets were substantially worse than the year before, largely due to the cluster exhibiting a double peak (Fig 3 C and D). The average single-peak line through the slightly diffuse TS data performed slightly better than did a similar line through the more sharply peaked PA data. We note that our data and models do not distinguish between subtypes of influenza A and that both H1N1 and H3N2 were circulating during similar periods in this season.

**Figure 3.**
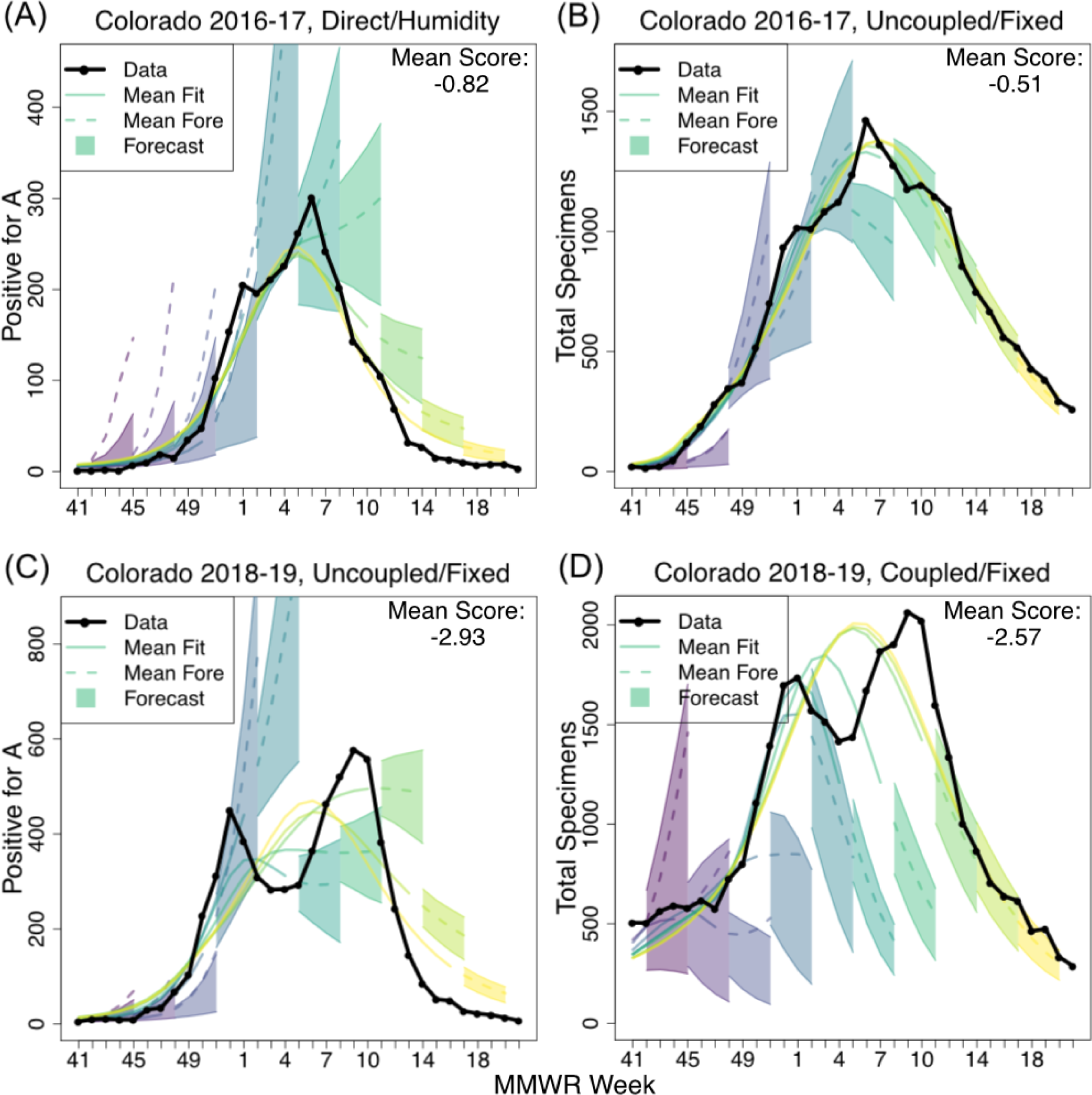
Illustrative high-scoring and lower-scoring forecasts for the Colorado cluster. Here the incidence data is shown in black and shaded cones depict the central 50% forecast windows for 1-4 week ahead. Forecasts were generated for every week, but only every third forecast is shown here for clarity. Colour progression from left to right indicates time of forecast. Solid lines show fit to data at the point the forecast was assumed to have been made. Dashed lines show forecasts beyond the time at which data were assumed to be available. Mean score is the average score across MMWR forecasted weeks and 1 to 4 weeks ahead targets.

Among the mechanistic models, coupled models generally performed better than uncoupled models or models that fit the cluster incidence directly (Fig 2). Also, the accuracy of mechanistic models relative to non-mechanistic average-based models compared for TS and PA data suggests that mechanistic models are able to capture more information from the PA data. For the TS data, the 5-week rolling historical average model outperformed the best mechanistic models for all time scales from 2-to 10-weeks. However, for PA data, even at a 10-week lead time, the best mechanistic model substantially outperformed the 5-week average historical model. We note that there were few years available to the historical models in these data, therefore the performance of historical-average models would likely improve over time as the training data set grows.

Patterns in the posterior densities for key mechanistic parameters also suggest that when the models are fit to the PA data they are capturing more of the underlying biology (Fig 4). The posterior median for the basic reproductive number *R*_*0*_ was consistent from year to year and location to location when fit to the PA data. For coupled models, cluster *R*_*0*_s were higher (mean 1.321; 95% CI [1.317, 1.324]) than for uncoupled models (mean 1.176; 95% CI [1.174, 1.177]). Given that transmissibility is reflected by *R*_*0*_*-1*, this difference is substantial and likely reflects the need for uncoupled models to have longer durations (resulting from lower *R*_*0*_) because the overall cluster incidence was not driven by epidemics in subpopulations taking off at different times. This theory is supported by an inverse relationship for the estimated proportion of infections which result in clinical cases *p*_*C*_. Estimates of *p*_*C*_ were lower for coupled models than for uncoupled models because subpopulations with higher *R*_*0*_s require fewer of their infections to result in cases to achieve the same level of incidence as those from the uncoupled model. Despite the coupled model outperforming the uncoupled model, estimates for the parameters that determined the strength of coupling were not well constrained when fit to the data (Fig S5).

**Figure 4.**
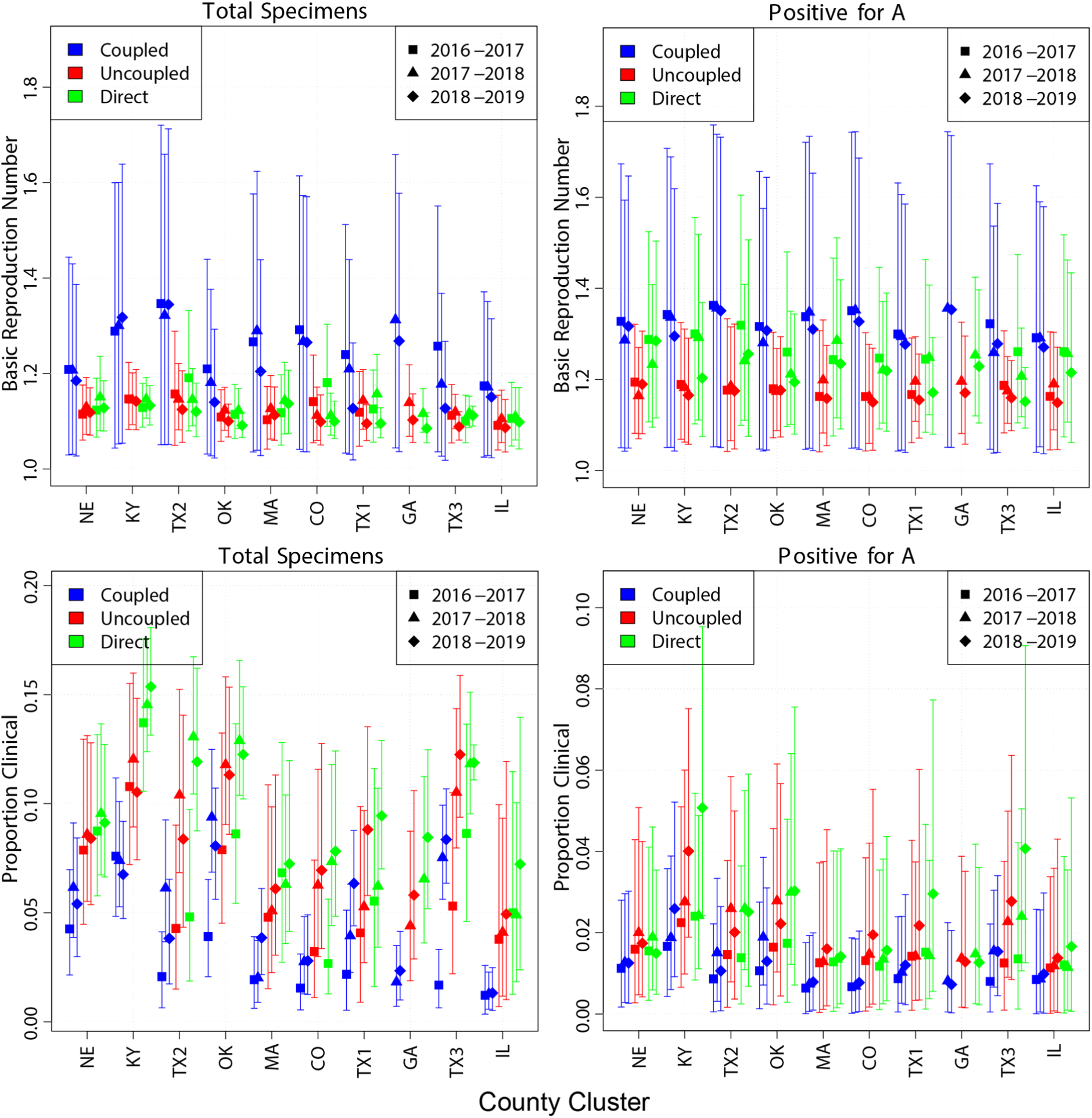
Posterior parameter densities by season, cluster and model type. Bars show central 95% of posterior credibility interval with symbol showing posterior mean. Colour indicates model type and symbol shape indicates season. Values for each cluster are grouped together as per x-axis tick labels.

## Discussion

We have shown that for a large set of mechanistic models, using a skill scoring system directly analogous to current best practise, cluster-level influenza incidence data based on type-confirmed test results are substantially better forecast than are data based only on the volume of respiratory samples being tested. We have also shown a close correlation between the number of respiratory samples being taken in each cluster and the pattern of influenza-like illness reported to the national surveillance system for the geographical region enclosing each cluster. Among these mechanistic models, those that fit to subpopulation county data in order to forecast cluster patterns are more accurate than those that do not. Also, the relative difference between the mechanistic models and simple historical average models is much greater when forecasting the type-specific data than when forecasting total samples.

A limitation of our study is that we compared only forecasts from our suite of mechanistic and historical average models against the two datasets. Therefore, it is possible that other approaches that we did not try may have been able to forecast the TS data more effectively than the PA data. However, the difference in our ability to forecast the two types of data was considerable and significantly greater than the difference in performance between our approach and other high performing participants in the CDC challenge study, which is measured on a similar scale (7). Also, intuitively, the shape of epidemics in the PA data was more consistent with a short-generation-time infectious disease epidemic: the degree of noise prior to the start of the season was low and the epidemic itself was more peaked, as measured by width at half maximum, suggesting that the substantial improvement in forecastability using a mechanistic approach is consistent with the PA data being much more representative of the underlying biological process.

A previous study has compared the forecastability of pathogen-specific data and found evidence of slightly increased forecastability of local type- and subtype-specific data, albeit using different accuracy metrics (12). However, the virological data available to the prior study represented a considerably higher spatial units than the separate ILI data. More local sub-type epidemics were inferred by combining regional laboratory-confirmed positive proportions with Google Flu Trends (16) estimates of state and municipality ILI data, implicitly assuming that the temporal distribution of types and subtypes was uniform for large geographical regions. We suggest that aggregated positive tests at the geographical and temporal scales of analysis contain maximal information that is not present in similar inferred data streams.

Our results suggest a number of additional refinements to our forecasting workflow that may further improve model forecast accuracy. We assumed that each cluster consisted only of the sub-populations for which data were available. It may be that the inclusion of epidemiological ‘dark matter’ -- the intervening and surrounding populations that were not observed -- could increase accuracy. This would substantially increase the computational complexity of the approach, but could potentially fill-in surveillance gaps at the same time as increasing the accuracy and robustness of the forecasts. Also, we have treated these as weekly data, which reflects their current reporting pattern. However, accuracy for now-casting and short term forecasting could be improved substantially by looking at daily incidence which are available from this surveillance network. Although some work would be required to make a day-of-week adjustment (17), accuracy of 1-week ahead and 2-week ahead forecasts would likely increase substantially over and above the gains described here.

Public health authorities may wish to consider the routine use of type- and subtype-specific point-of-care data for influenza surveillance in addition to traditional ILI surveillance to enable more accurate local influenza forecasts during seasonal and pandemic periods. Our results were for 10 geographical clusters based only on three seasons worth of data (so a maximum of two training years). Very soon, the network of machines from which these data were obtained and other similar networks will have much more complete geographical and temporal coverage (18). Similar networks are being established in many other populations. While solely laboratory-based virological data have historically suffered from dramatic changes in testing volumes at the start of a pandemic, there is no reason to believe that the same changes will occur in these largely non-hospital point-of-care machines: or that any such changes should they occur would be worse than those that occur in influenza-like illness surveillance networks.

The accuracy of retrospective forecasts presented here extends prior understanding of local dynamics of the transmission of influenza, and suggests plausible novel public health strategies. Other studies have shown that aggregating local forecasts can improve accuracy over direct forecasts for larger geographical units, but have not focussed on the potential accuracy at small spatial scales (10, 19). Also, more descriptive ecological analyses have demonstrated the potential for improved local forecasts by characterizing how humidity and population density are correlated with average properties of epidemic curves (20). Building on these prior studies, the smaller spatial unit and temporal accuracy of the results presented here could be used as a public health intervention to give individuals the opportunity to modify their behaviour over short periods of time, based on very local information. In particular, people at high risk of severe disease may choose to reduce their social mixing during short periods where the risk of infection locally can be accurately predicted to be high.

## Materials and Methods

### Deployment of test machines and capture of test data

Our data are gathered from a near-real time network of benchtop Sofia and Sofia 2 POC diagnostic machines (Quidel, San Diego, USA). At the start of the study period in July 2016, the network consistent of 1676 machines located in physician offices (880), hospitals (257), urgent care clinics (427) and other/not reported (112) settings. At the end of the study period the network had grown to 12345 machines with 5,736 in physician offices, 1,206 in hospitals, 1,194 in urgent care clinics, and 4,209 in other/not reported settings. The standard kit tests for influenza A and B. Data are automatically transmitted to Quidel. Reporting cadence can vary depending on device connectivity, but most devices report results within a day of testing.

### Selection of counties and clusters

Individual strain tests were sorted into weekly totals for each county represented in the dataset. Of the approximately 3240 counties and county equivalents in the United States, 1258 are represented in this dataset. However, to omit sparse data streams, counties that did not report for at least 40 weeks in both the 2016-17 and 2017-2018 seasons were discarded. Next, counties with less than 250 specimens tested in any single season of 2016-17, 2017-18, or 2018-2019 were discarded. These requirements reduced the number of considered counties from 1258 to 136. This county filtering process was based on data reported up to and including week 3 of 2019.

To identify localized groups within the 136 qualifying counties, we applied a k-means clustering algorithm for the number of clusters *k*. Cluster distances were based on county population centroids *c* (lat/lon). For a single cluster *a*_*j*_ with *N* counties and geographic cluster centroid *b*_*j*_, we define the intra-cluster distance as the sum of squares

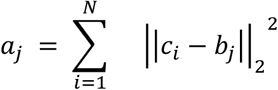

where *c*_*i*_ is the centroid of the *i-*th county of cluster *j*. The k-means algorithm was applied while varying *k* over the interval *[2, 60]*. The total intra-cluster sum of squares (over all clusters) stopped decreasing at approximately *k=38*. So we chose *k=38*. Smaller, sparsely spaced clusters were removed by requiring *N* ≥ 4 and *a*_*j*_ /*N* ≤ 0.5.

The one exception to these rules is the Atlanta cluster. Most Atlanta counties do not pass the incidence minimums for the 2016-2017 season. However, county coverage for 2017-18 and 2018-19 is the best of any large metropolitan area in the United States, so Altlanta was included for these seasons.

This process resulted in 10 county clusters, covering 57 counties as described in Table S2.

### Mechanistic modelling framework

The following sections offer a summary of the model framework, covariate data collection, and fitting procedure used for this work. For a more complete technical discussion of these topics, see (10). These concepts were executed using the software package Dynamics of Interacting Community Epidemics (DICE). DICE is a publicly available R-package available for download with documentation and examples (see Data Availability).

The foundation of our model is the common Susceptible-Infectious-Recovered (SIR) set of equations (21). We include terms to modulate the force of infection for specific humidity and county coupling (10). The probability of inter-county transmission is evaluated using a distance-based mobility kernel. Here, the distance between counties is calculated as the Euclidean distance between counties’ population centroids.

The spatial aspect of the model is characterized in three ways: direct, uncoupled, and coupled. The self-describing ‘direct’ fit matches the model directly to cluster data. When applying the uncoupled model to a cluster, each constituent county is fit independently. The individual county forecasts are then aggregated to produce a cluster forecast. The coupled approach simultaneously fits the coupled model to all constituent counties.

Specific humidity data was taken from the National Centers for Environmental Prediction (NCEP)/National Center for Atmospheric Research (NCAR) Reanalysis-2 project (22). This provides climate model evaluations 1979-present on a 1.875° spatial grid. The reanalysis-2 spatial grid is layered with GADM (the Database of Global Administrative Areas) county definitions (23) and the SocioEconomic Data and Applications Center (SEDAC) population densities (24) to produce weekly population-weighted averages for specific humidity in each county.

### Fitting the models

Here we define incidence as the population count that was ILI symptomatic (TS) or influenza infectious (PA) *and* presented to a reporting clinic for testing. Model equations are applied to the full population, so incidence is returned from the model as a proportion of the new infectious. This proportional scaling accounts for both the portion of the population covered by reporting clinics and the proportion of the infectious population that sought medical evaluation/treatment.

For each fit data series, we calculate the Log-Likelihood (LLK) that the data is a Poisson expression of model values. We then use a Metropolis-Hastings type Markov Chain Monte Carlo (MCMC) process (27) with adaptive step size to map the likelihood in parameter space. When the coupled model is used, LLK of the cluster is calculated as a population-weighted average of the individual county LLKs.

### Forecast Scoring

For each forecast target, our model produces a distribution of outcomes. This distribution is binned for evaluation based on the scoring system used by the CDC Influenza Challenge (8) for the 2016-17, 2017-18 and 2018-19 seasons. The maximum range is separated into 130 equally sized bins with a 131st bin for values over the maximum previously observed. We re-scaled the bins for total number of tests and the number of tests positive for influenza A so that all three metrics peak in the same bin (Figure S1). Forecast probabilities for the observed bin (red shading) as well as the 5 pre- and pro-ceeding bins (green shading) are summed to produce forecast skill. Forecast score is the natural log of skill, with scores below −10 set to −10. Note that due to (28) and the response (29), starting with the 2019-20 influenza season, the CDC Influenza challenge will define forecast skill as only the probability in the observed bin. Here we retain the 11-bin skill window for consistency with matching Influenza Challenge seasons.

### Null Models

For a given season, cluster, week, and data metric, Null models were evaluated using data from the same cluster, metric, and week, but different seasons. The most basic Null model ‘N.pt’ is a point forecast calculated as the mean of the Null data. The ‘N.d’ model is a distribution fit to the same data. For intensity targets we use a log normal distribution and for timing targets a normal distribution. However, both of these models suffer from the same problem: due to the small number of seasons in the dataset, the model is typically being fit to only 2 data points. The third Null model ‘N.d5’ was designed to mitigate this problem. For a given forecast week, ‘N.d5’ uses the historic data from a 5-week window including the forecast week as well as the 2 pre- and pro-ceeding weeks to fit a log normal distribution. Ex. for epidemic week 4 of 2017, the data from epidemic weeks 2, 3, 4, 5, and 6 for years 2018 and 2019 are fit to a log normal. This distribution then gets to binned to serve as the N.d5 model forecast.

### Data availability statement

All weekly incidence data for the smallest spatial unit (counties) are available immediately from the authors for non-commercial use in a format that can be used immediately with the forecasting tools used to generate these results. The forecasting tools are available with documentation and examples from https://github.com/predsci/DICE_quidel_manuscript. [Note to reviewers and editors - this URL is for review only. At final publication, this will be changed to the standard public URL for DICE].

## Data Availability

All weekly incidence data for the smallest spatial unit (counties) are available immediately from the authors for non-commercial use in a format that can be used immediately with the forecasting tools used to generate these results. The forecasting tools are available with documentation and examples from https://github.com/predsci/DICE_quidel_manuscript.

## Acknowledgements

We thank John Timerius (Quidel Inc) for the provision of these data and for helpful discussions.

## Supporting Information

**Figure S1.**
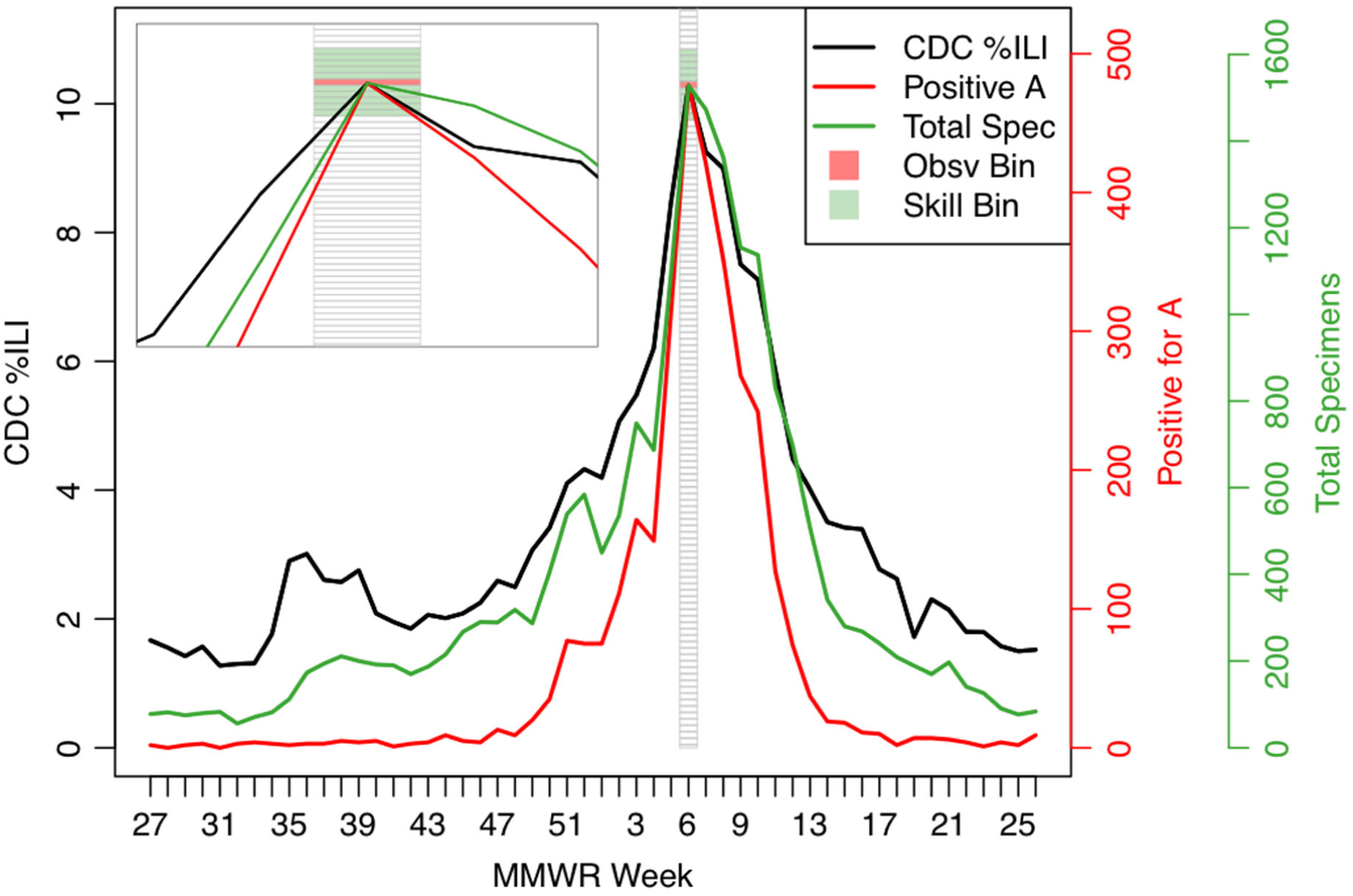
Similarity between total samples time series and influenza-like illness data and bin-scoring design, illustrated for 2016-2017 season in the San Antonio / Austin area cluster. Bins were scaled to each metric such that all peaks fall in the same bin. The skillful bins for the peak week are shown in green shading. Forecast probabilities in these bins, in addition to the observed bin (red shading), are summed to calculate skill. The Centers for Disease Control and Prevention percentage ILI (CDC %ILI) is for the enclosing CDC region (state of Texas).

**Figure S2.**
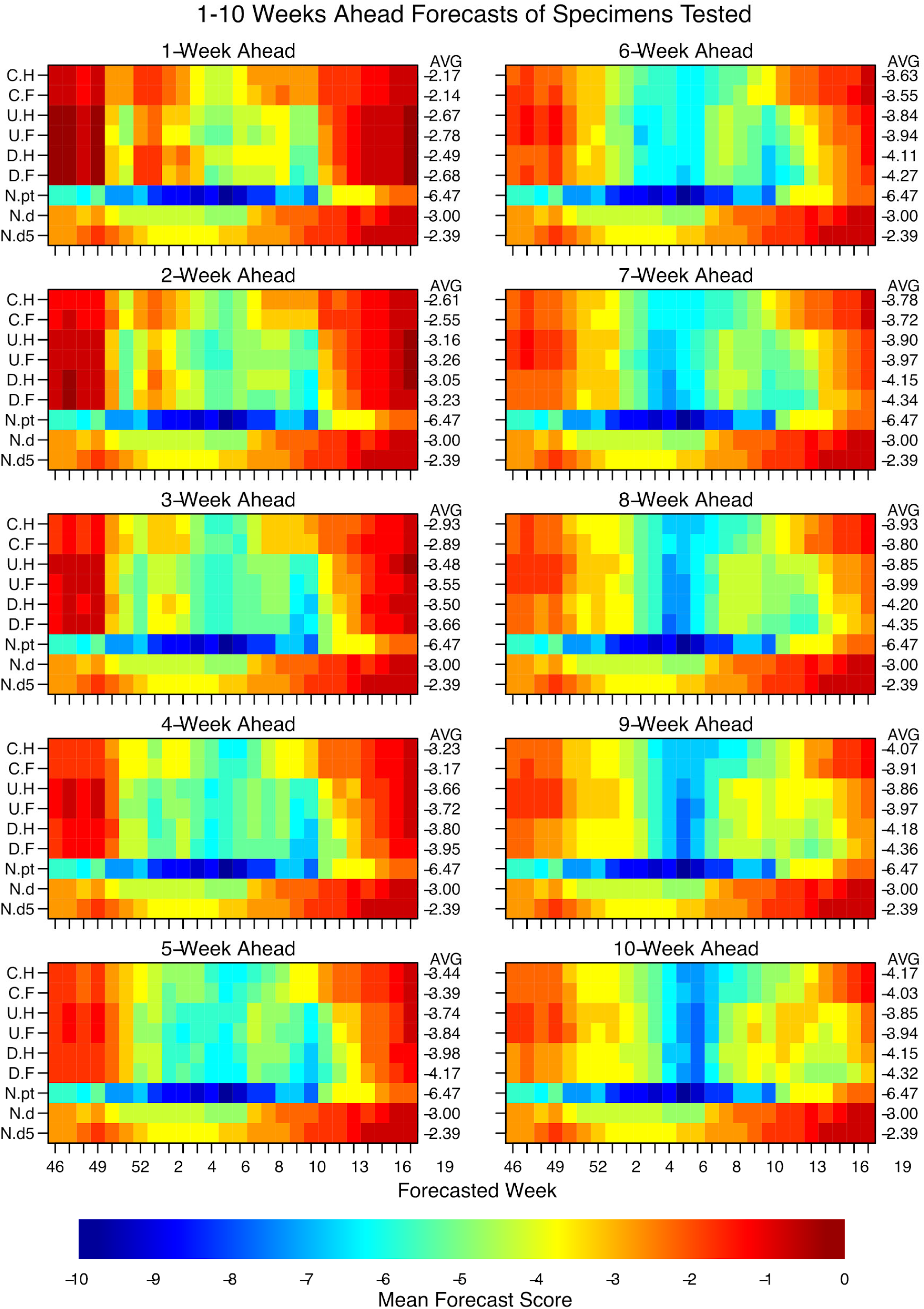
Mean scores for Total Specimens forecasts 1-10 weeks ahead. Pixel colour shows forecast score (see main text) for a given observation week averaged across clusters and seasons, e.g. the 4-week ahead forecast for week 48 was made using data only upto week 44. Averages across all weeks for a given model are printed on the RHS of each row of pixels. Model type is shown on LHS y-axis tick labels: C.H, coupled model with humidity modulated contact rate; C.F, coupled model with fixed contact rate; U.H, uncoupled with humidity; U.F, uncoupled with fixed contact rate; D.H, model directly fitted to cluster with humidity term; D.F, model directly fitted to cluster with fixed contact rate; N.pt, null model made from simple model of that week for all other seasons; N.d, null model made from fitting a log normal to all observations for that week from other years; and N.d5, null model made from fitting log-normal to the observation week, two weeks prior and two weeks following for all other years (see main text). Models are ordered approximately from least complex on the bottom rows to most complex on the top row.

**Figure S3.**
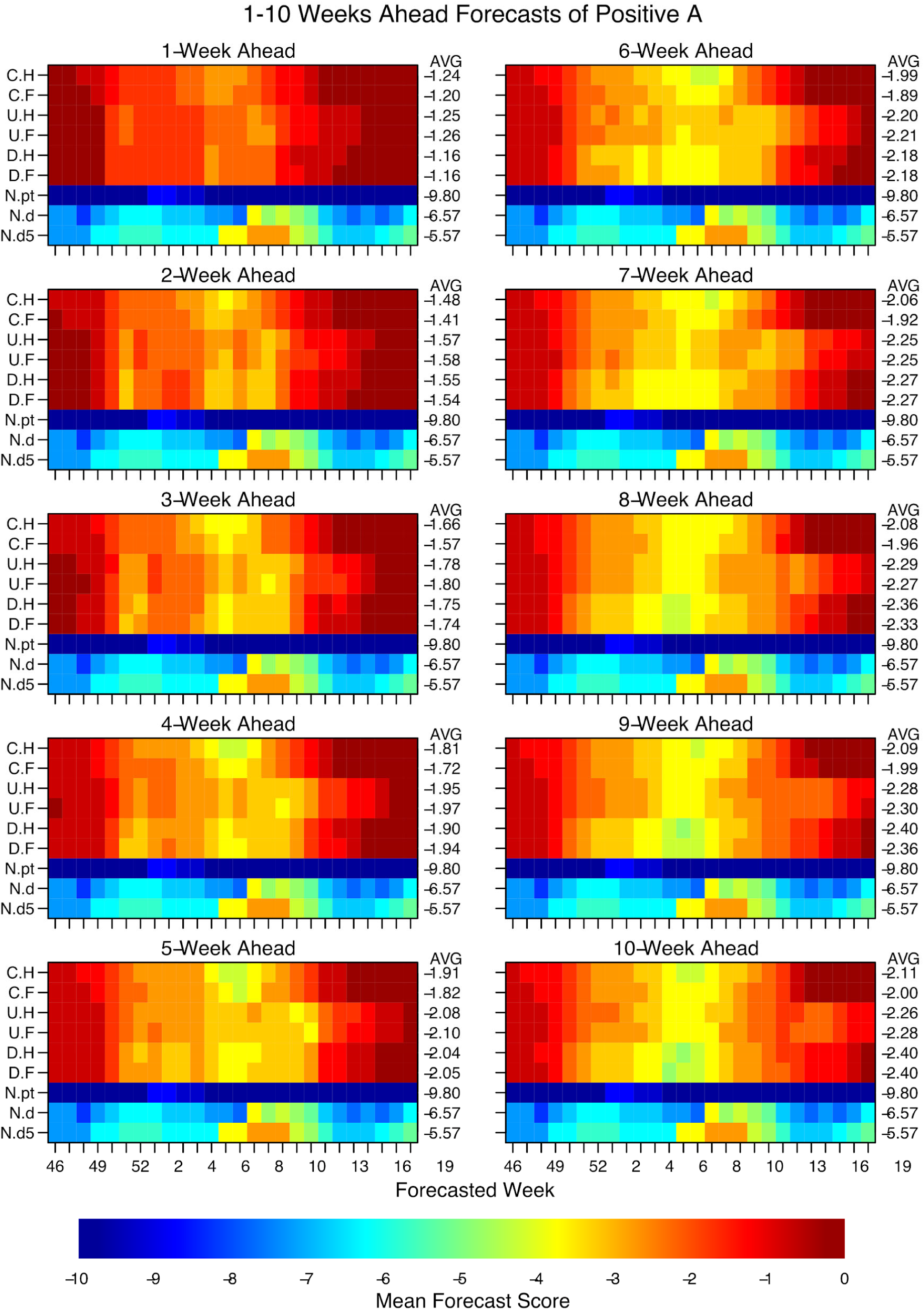
Mean scores for Positive-for-A forecasts 1-10 weeks ahead. Pixel colour shows forecast score (see main text) for a given observation week averaged across clusters and seasons, e.g. the 4-week ahead forecast for week 48 was made using data only upto week 44. Averages across all weeks for a given model are printed on the RHS of each row of pixels. Model type is shown on LHS y-axis tick labels: C.H, coupled model with humidity modulated contact rate; C.F, coupled model with fixed contact rate; U.H, uncoupled with humidity; U.F, uncoupled with fixed contact rate; D.H, model directly fitted to cluster with humidity term; D.F, model directly fitted to cluster with fixed contact rate; N.pt, null model made from simple model of that week for all other seasons; N.d, null model made from fitting a log normal to all observations for that week from other years; and N.d5, null model made from fitting log-normal to the observation week, two weeks prior and two weeks following for all other years (see main text). Models are ordered approximately from least complex on the bottom rows to most complex on the top row.

**Figure S4.**
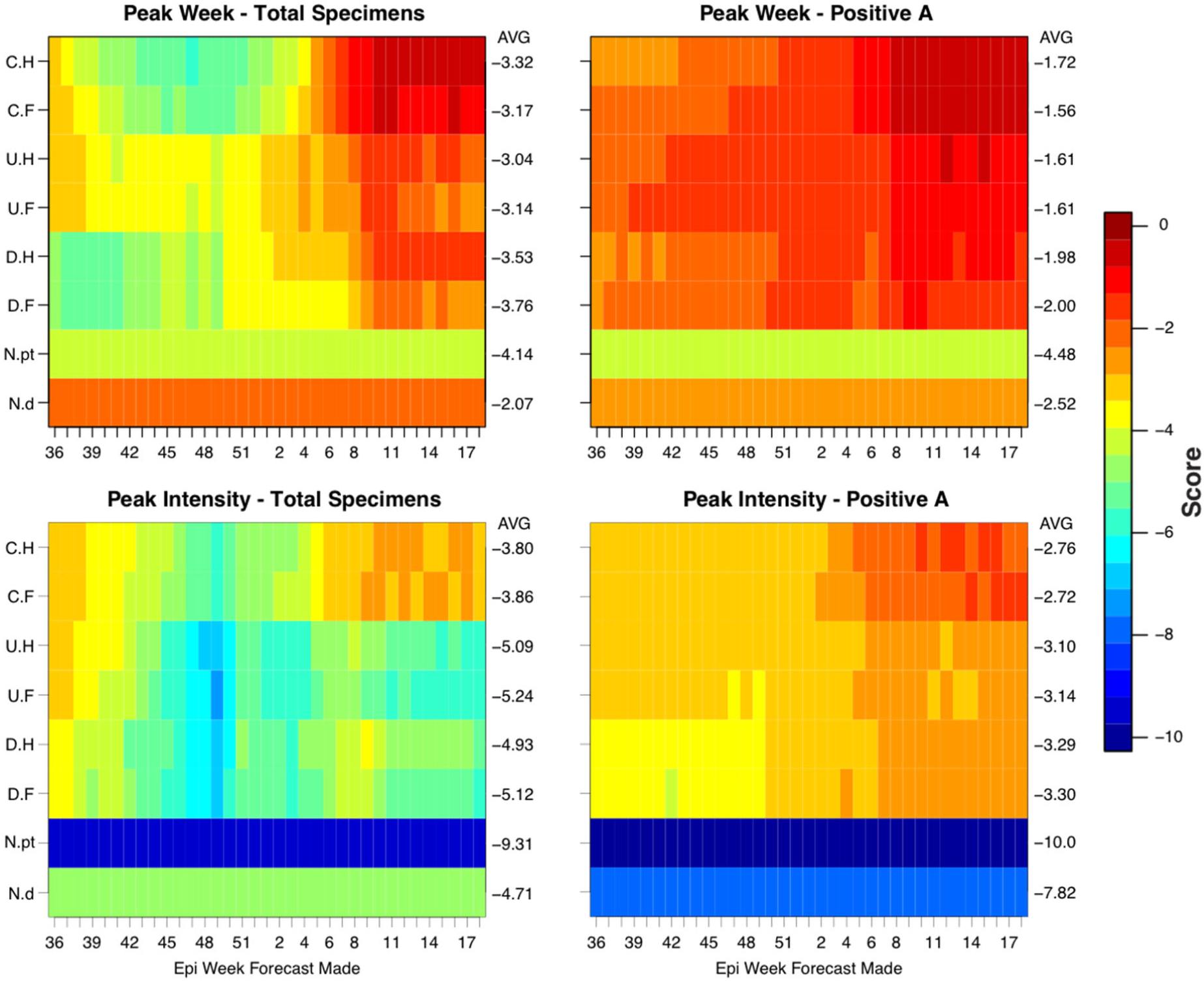
Peak target mean forecast scores for total specimens and positive for A. The panels are divided by target (top - Peak Week, bottom - Peak Intensity) and data metric (left - total specimens tested, right - specimens positive for A). Forecast scores for each square are averaged across all clusters and all seasons. The mean value of each row appears in the AVG column on the right. The first column of model abbreviations is interpreted: C-coupled, U-uncoupled, D-direct, N-Null. The second column of abbreviation is: H-humidity modulated contact rate, F-fixed contact rate, pt-point forecast, d-log normal distribution.

**Figure S5.**
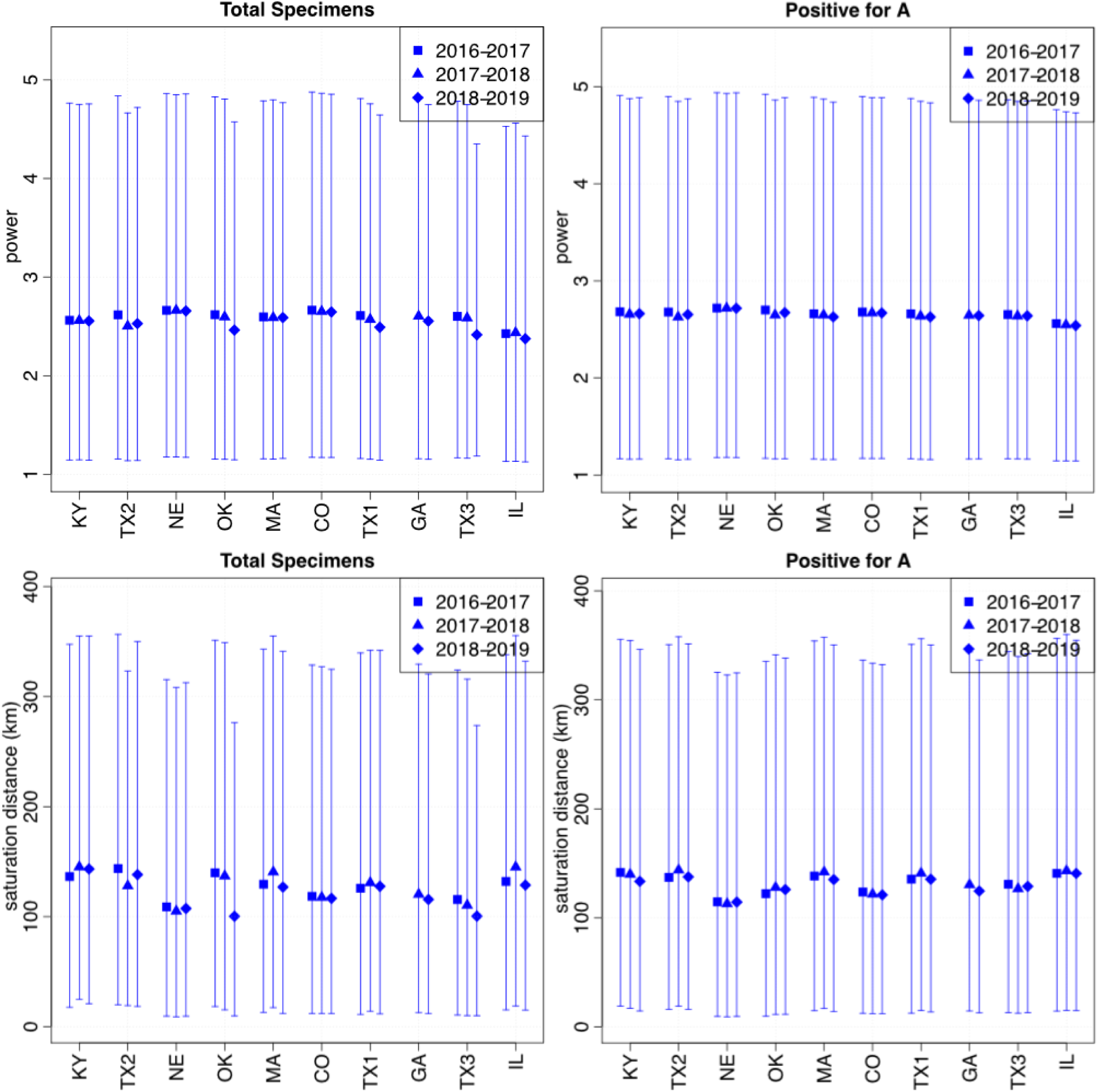
Coupling parameter mean and central 80% interval. Results were compiled from MCMC chains across all models and forecast weeks for a given cluster and season. Coupling is modeled with the offset power law relationship described in (10) using free parameters for saturation distance and power. Briefly, the probability of coupling occurring across space is determined by a function with a plateau followed by a power law drop-off. Saturation distance is the width of the plateau and ‘power’ is the power coefficient of the drop-off.

**Table S1.** Pearson correlations between data metrics. Correlations are calculated for each cluster and season, and comparing total specimens, positive for, A and CDC %ILI. From the resulting distributions, we have extracted the median, 20%, and 80% quantiles.

| Metrics Compared | Median | 20% | 80% |
| --- | --- | --- | --- |
| CDC %ILI v Total Spec | 0.955 | 0.928 | 0.973 |
| CDC %ILI v Positive A | 0.924 | 0.860 | 0.945 |
| Total Spec v Pos A | 0.937 | 0.911 | 0.958 |

**Table S2.** County cluster information.

| Abbv | Title Short | Title Long | CDC Parent | Counties |
| --- | --- | --- | --- | --- |
| CO | Colorado | Colorado Front Range | Colorado | Adams, Arapahoe, Boulder, Douglas, El Paso, Jefferson, Pueblo, Weld |
| GA | Atlanta | Atlanta Metropolitan Area - GA | Georgia | Bartow, Carroll, Clayton, Cobb, Coweta, Forsyth, Fulton, Gwinnett, Hall, Henry, Paulding |
| KY | Kentucky | Central Kentucky | Kentucky | Hardin, Mercer, Nelson, Warren |
| MA | Massachusetts | Massachusetts | Massachusetts | Bristol, Essex, Norfolk, Plymouth, Worcester |
| OK | Oklahoma City | Oklahoma City - OK | Oklahoma | Cleveland, Comanche, McClain, Oklahoma |
| IL | Chicago | Chicago Area - IL,IN,WI | Region5 | Cook, Lake, Dane, Outagamie, Winnebago |
| NE | Omaha | Omaha - NE,IA | Region7 | Harrison, Pottawattamie, Douglas, Sarpy |
| TX1 | South TX | San Antonio/Austin - TX | Texas | Bexar, Comal, Travis, Williamson |
| TX2 | Texarkana | Texarkana - TX,AR | Texas | Miller, Bowie, Gregg, Harrison, Hopkins |
| TX3 | Dallas | Dallas/Ft Worth - TX | Texas | Collin, Dallas, Ellis, McLennan, Somervell, Tarrant, Wise |

